# Performance on the Balloon Analogue Risk Task and Anticipatory Response Inhibition Task is Associated with Severity of Impulse Control Behaviours in People with Parkinson’s Disease

**DOI:** 10.1101/2022.10.20.22281277

**Authors:** Alison Hall, Matthew Weightman, Ned Jenkinson, Hayley J MacDonald

**Author notes:** Corresponding Author: Hayley MacDonald, Department of Biological and Medical Psychology, University of Bergen, Bergen, Norway.

## Abstract

**Introduction:** Dopamine agonist medication is one of the largest risk factors for development of problematic impulse control behaviours (ICBs) in people with Parkinson’s disease. The present study investigated the potential of dopamine gene profiling and individual performance on impulse control tasks to explain ICB severity.

**Methods:** Clinical, genetic and task performance data were entered into a mixed-effects linear regression model for people with Parkinson’s disease taking (n = 50) or not taking (n = 25) dopamine agonist medication. Severity of ICBs was captured via the Questionnaire for Impulsive-compulsive disorders in Parkinson’s disease Rating Scale. A cumulative dopamine genetic risk score (DGRS) was calculated for each participant from variance in five dopamine-regulating genes. Objective measures of impulsive action and impulsive choice were measured on the Anticipatory Response Inhibition Task and Balloon Analogue Risk Task, respectively.

**Results:** For participants on dopamine agonist medication, task performance reflecting greater impulsive choice (p = .014), and to a trend level greater impulsive action (p = .056), as well as a longer history of DA medication (p < .001) all predicted increased ICB severity. DGRS however, did not predict ICB severity (p = .708). No variables could explain ICB severity in the non-agonist group.

**Conclusions:** Our task-derived measures of impulse control have the potential to predict ICB severity in people with Parkinson’s and warrant further investigation to determine whether they can be used to monitor ICB changes over time. The DGRS appears better suited to predicting the incidence, rather than severity, of ICBs on agonist medication.

## INTRODUCTION

Problematic impulse control behaviours (ICBs), incorporating impulse control disorders and other related behaviours, can develop in Parkinson’s disease (PD) patients. These behaviours often manifest as compulsive gambling, binge eating, hypersexuality, compulsive shopping, punding, hobbyism and compulsive medication use (Weintraub 2008). ICBs are routinely identified using the questionnaire for impulsive-compulsive disorders in Parkinson’s disease (QUIP) and further clinically diagnosed during an interview (Weintraub et al. 2009; Papay et al. 2011; Weintraub et al. 2012; Probst et al. 2014; Krieger et al. 2017; Marques et al. 2019; Takahashi et al. 2022). The Questionnaire for Impulsive-compulsive disorders in Parkinson’s disease short (QUIP-S) and QUIP rating scale (QUIP-RS) are two widely used self-report versions of this questionnaire. The QUIP-S involves only 13 questions with ‘yes’ or ‘no’ answers (Weintraub et al. 2009; Krieger et al. 2017), whereas the QUIP-RS includes 28 questions which are answered via a frequency rating scale with five different options and the final score is equated with ICB severity (Weintraub et al., 2012; Probst et al., 2014; Marques et al., 2019; Takahashi et al., 2022). The QUIP-RS offers a larger range of scores covering the same behaviours in more depth, which suggests the resultant ICB frequency (i.e., severity) score is capable of being a more sensitive measure of impulsivity, including changes over time (Marques et al. 2019), compared to ICB incidence from the QUIP-S (Weintraub et al. 2012; Probst et al. 2014). The Barratt Impulsiveness Scale (BIS) is also a self-report questionnaire that measures impulsivity but as a trait or personality construct (Stanford et al., 2009), rather than a diagnostic tool for pathological ICBs directly. Nevertheless, ICBs in PD determined by the QUIP-S (Marin-Lahoz et al. 2018) and QUIP-RS (Takahashi et al. 2022) are associated with higher impulsivity on the BIS. One previous investigation determined a positive correlation between total QUIP-RS score and BIS score (Goerlich-Dobre et al. 2014), highlighting the potential adjunct use of the BIS in ICB diagnosis.

One of the most significant risk factors for ICBs in PD is dopamine agonist (DA) medication, where 14-40% of patients taking this form of dopamine replacement therapy develop destructive ICBs (Bastiaens et al. 2013; Kraemmer et al. 2016; Erga et al. 2018). Clinically prescribed DAs predominantly act upon D2/D3 receptors (Gasser et al. 2015; Seeman et al. 2015), which are abundant in regions of the mesocorticolimbic (MCL) system (Ko et al. 2013; Seeman, 2015). The MCL system is largely responsible for impulse control and is relatively spared during the early, unmedicated stages of PD (Cools, 2006; Weintraub, 2008; Smith et al. 2016; Caminiti et al. 2017; Claassen et. al. 2017; Gatto & Aldinio, 2019), compared to the decrease of dopamine in the nigrostriatal system (Dauer & Przedborski, 2003; Weintraub, 2008; Vaillancourt et al. 2013). It is therefore possible that the addition of DA medication causes a tonic hyperdopaminergic state in the MCL network, which hinders phasic dopamine modulation, and subsequent problems with impulsivity (Weintraub, 2008; Sinha, Manohar & Husain, 2013; Vaillancourt et al. 2013; Gatto & Aldinio, 2019; Meder et al. 2019). This state has been termed the overdose-hypothesis (Cools et al. 2001a; Vaillancourt et al. 2013; Ruitenberg et al. 2021). Moreover, increases in DA dose and the use of DA medication over time are often associated with ICBs in PD, due to higher concentrations of dopamine activating D2 receptors to a greater extent compared to lower concentrations (Trantham-Davidson et al. 2004). The working hypothesis being that increased and/or prolonged receptor activation may reduce D2 auto-receptor sensitivity (Gasser, Wichmann & DeLong, 2015), leading to a blunted post-synaptic D2-mediated inhibitory effect, increased overall dopamine release and resultant impulsive behaviour (Ray et al., 2012; Ford, 2014). The two possible mechanisms of effect are not mutually exclusive, and may well act in concert, though both offer explanations as to why DA medication leads to dysfunctional levels of dopamine and ICB development in some patients.

Another factor which can influence ICB development is genetic. Previous literature has identified specific genetic polymorphisms associated with ICBs in PD patients, either individually (Lee et al. 2009; Kraemmer et al. 2016; Erga et al. 2018) or collectively as a very large polygenic risk score (Ihle et al. 2020, Faouzi et al. 2021). The first dopaminergic genetic score quantifying the influence of a small number of genes was developed by Nikolova and colleagues (2011). This method was subsequently expanded by Pearson-Fuhrhop and colleagues (2013, 2014) to produce a polygenic dopamine genetic risk score (DGRS) incorporating five specific genes selected a-priori for each being known to modify dopamine signalling within MCL regions (Vriend et al. 2014; Smith, Xie & Weintraub, 2016; Caminiti et al. 2017) and influence impulse control (Congdon et al. 2009; Lee et al. 2009; Vriend et al. 2014; Abidin et al. 2015; Smith, Xie & Weintraub, 2016; Erga et al. 2018). These genes include: DRD1 rs4532, DRD2 rs1800497, DRD3 rs6280 (encoding D1, D2, D3 receptors, respectively), catechol-O-methyltransferase (COMT) rs4680 and dopamine transporter (DAT) rs28363170. The quantitative aspect of the DGRS weights the influence of each polymorphism on widespread tonic dopamine neurotransmission, where a higher score is equal to higher dopamine neurotransmission. It stands to reason that a PD patient’s genetically determined levels of MCL dopamine neurotransmission will affect how they respond, and whether they develop ICBs, when dopamine tone is further increased with DA medication. Indeed, our previous work utilising the DGRS for the first time in PD (Hall et al. 2021) demonstrated that patients with a low DGRS had more ICBs identified via the QUIP-S, which decreased with time on DA medication. Conversely, patients with a higher DGRS had fewer ICBs, but this number increased with time on DA medication. We were unable to discern whether increasing dosage over time or time of exposure to DA medication per se were causing these changes in ICBs.

MacDonald and colleagues (2016) were first to use the DGRS to explain objective measures of behavioural impulsivity in a non-PD population. These objective measures were stop signal reaction time (SSRT) from the Anticipatory Response Inhibition Task (ARIT) for impulsive action, and decision making following negative reinforcement on the Balloon Analogue Risk Task (BART) for impulsive choice. They concluded that the administration of DA medication in healthy adults improved task measures of impulsive action and choice for those with a lower DGRS and worsened them for participants with a high DGRS. Previous literature has identified no change in impulsive behaviour for PD ICB patients after a loss on the BART, compared to non ICB patients who reduced their impulsive behaviour (Martini et al. 2018). Either shorter or no difference in SSRT has been found for ICB vs no ICB PD patients in the Stop Signal Task (Claassen et al. 2015; Ricciardi et al. 2017; Vriend et al. 2018; Hlavata et al. 2020). The ARIT and our specific measure of negative reinforcement in the BART have yet to be investigated in a PD cohort in the context of ICBs.

Thus, the primary aim of the present study was to investigate whether task-derived measures of impulsivity and a DGRS were associated with clinically identified impulsive behaviours via the QUIP-RS in a sample of PD patients taking DA medication, and whether there were any interactions between these variables. We hypothesised that patients with a low DGRS would display worse impulsivity (i.e., higher SSRT and a negative reinforcement value further from zero) and higher ICB frequency. Whereas those with a high DGRS would exhibit better impulsivity on the tasks and lower ICB frequency. A secondary aim was to determine if DA medication dosage or time of exposure to DA medication could predict ICB frequency, following our previous results (Hall et al. 2021). We also hypothesised that both DA medication dosage and time on DA medication would be higher for patients reporting a greater frequency of ICBs. When accounting for the influence of an individual’s genetic profile, we hypothesised that for patients with a low DGRS, longer exposure to DA medication would result in a reduction in ICBs over time. In contrast, patients with a high DGRS were expected to show increasing ICB frequency with increasing time on DA medication. We did not expect to find any comparable results for patients taking dopamine medication which did not include DAs. Finally, we wanted to examine any relationship between clinically identified ICBs and subjective trait impulsivity via the BIS.

## MATERIALS AND METHODS

### Participants

One hundred participants with PD were recruited for the current study; 70 were taking DA medication and the remaining 30 were taking dopamine medication not including agonists. Participants were included in the study if they were between the ages of 40-80, had no history of neurological illness other than PD and had normal or corrected-to normal vision. All demographic, clinical, questionnaire, behavioural and genetic data were collected remotely or online.

### Clinical Impulsivity

#### ICB incidence

The QUIP-short comprised of 13 ‘yes’ or ‘no’ questions regarding current impulse control behaviors lasting at least 4 weeks. Participants would receive a score of one for ‘yes’ and zero for ‘no’. Any score greater than zero confirmed the incidence of an ICB.

#### ICB frequency

The QUIP-RS measured the frequency of ICBs. The questionnaire included four questions in each of the following categories: gambling, sex, buying, eating, hobbyism, punding and PD medication. Participants responded to each question with a choice from a 5-point scale (0: never, 1: rarely, 2: sometimes, 3: often, 4: very often) which represented impulsivity in the past 4 weeks or any 4-week period in a designated time frame. Total scores were calculated between 0-112.

#### Trait Impulsivity

##### Barratt Impulsiveness Scale

A 4-point scale (1: rarely/never, 2: occasionally, 3: often, 4: almost always/always) questionnaire comprising of 30 questions about everyday behaviours assessing attentional, motor and non-planning trait impulsivity (Patton et al., 1995). A higher score reflects greater impulsivity. Two patients did not provide answers for 2 questions relating to the work environment as they were retired, and one patient did not answer one of the questions.Therefore, each participant’s result was normalised to a percentage where the score was divided by the total score possible from the number of questions answered and then multiplied by 100.

##### Impulsivity Task Performance

###### Anticipatory Response Inhibition Task (ARIT)

The ARIT was presented on a computer screen using custom code written in Inquisit 6 Lab (Version 6.5.1, Millisecond Software) and responses were made using a keyboard. Participants completed the task on their personal computers at home. Participants initially observed an instruction video and practised 20 Go and 9 Stop trials. Subsequently, they were required to complete 10 blocks of 40 experimental trials. The experimental trials consisted of 295 Go trials and 105 Stop trials in a randomised order.

For the experimental procedure, on each trial participants were presented with a screen containing two vertical white bars (Figure 1). The left bar was controlled with the ‘z’ key using the left index finger and the right bar was controlled with the ‘? /’ key using the right index finger. Every trial started with the participant holding down both keys which initiated a black bar rising within each of the white bars. Both black bars rose at equal rates and filled the white bars completely after 1000ms. The black bars intercepted a horizontal target line at 800ms. During Go trials, participants were required to intercept the horizontal target line with the rising bars by timing the removal of their fingers from both keys appropriately (successful releases were within 40ms above the target and 30ms below). Stop trials consisted of Non-Selective Stop Both (SB) trials and Partial Stop trials. During SB trials, participants were asked to keep both keys depressed when both bars stopped rising before reaching the target (Figure 1). Partial Stop trials comprised of Stop Left (SL) and Stop Right (SR) trials, where one bar stopped and the other continued rising. Here, participants were required to keep the key depressed corresponding to the bar that stopped rising and intercept the target line with the alternative bar by releasing the corresponding key (Figure 1). During Stop trials the bars initially stopped at 400ms for SB and 300ms for SL and SR. A staircase algorithm was utilised to generate a 50% success rate for each stop version. Following a successful Stop trial, the bar stop time increased by 25ms on the subsequent Stop trial but decreased by 25ms following an unsuccessful Stop trial. Stop signal reaction time from SB trials was calculated as the primary dependent measure using the integration method (Logan & Cowan, 1984; Verbruggen et al. 2019).

**Fig. 1.**
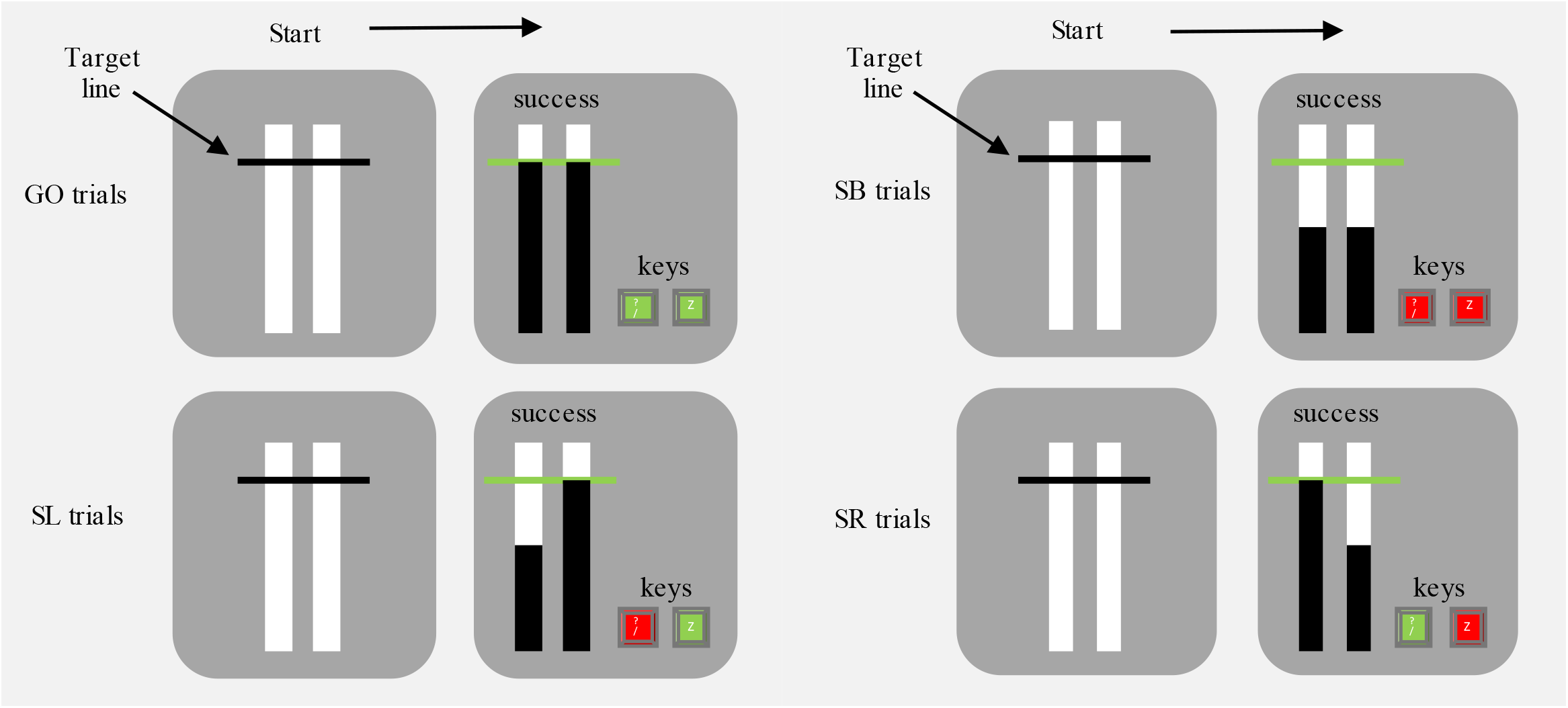
Visual display at the start of a trial (all left panels) and during a GO, SB (Non-Selective Stop Both), SL (Stop Left) and SR (Stop Right) trial in the Anticipatory Response Inhibition Task. Green keys represent successful release of the key at the target and red keys represent successful cancellation and keeping the key depressed. On successful Go trials, both keys are released at the target line. On successful SB trials, both keys are held down. On successful SL trials, the right key is released, and the left key is held down. On successful SR trials, the left key is released, and the right key is held down.

###### Balloon Analogue Risk Task (BART)

The BART was displayed on the participant’s personal computer screen using custom code written in Inquisit 6 Lab (Version 6.5.1, Millisecond Software) and responses were made using the mouse. Participants initially completed 5 practise trials and then 30 experimental trials.

The experimental procedure was as follows: at the beginning of each trial, participants were presented with a new balloon and two options: ‘Pump up the balloon’ or ‘collect £££’ (Figure 2). Participants could pump up the balloon, which incrementally increased potential earnings by £0.02 with each pump. If participants chose to collect their earnings, then the current trial would end, and the amount accumulated was added to the total winnings. However, the balloon could randomly explode on any pump and any potential earnings would be lost, followed by the end of the trial. Each trial started with a 1 in 85 probability of the balloon exploding. With every pump of the balloon, one number was randomly selected and removed without replacement from an 85-length array. When number one was selected, the balloon would pop and the trial would end with no monetary collection. The risk of balloon explosion therefore increased with each pump (1/84, 1/83 etc), but so did the potential monetary reward. The average number of pumps on a collection trial (i.e., when the number of pumps was not artificially constrained by a balloon burst) following a successful monetary collection (average collection pumps) and following a loss (balloon explosion) were calculated for each participant. The difference between these means normalised to pumps after a loss (losses cancel) reflected positive reinforcement and normalised to pumps after a win (wins cancel) reflected negative reinforcement (Mata et al., 2012; MacDonald et al., 2016). Proportions further from zero indicated a greater change in behaviour following either a positive or negative outcome. In this context, behaviour modification reflects a change in impulsivity. Negative reinforcement was the main dependent measure in this task, given previous results (MacDonald et al. 2016).

**Fig. 2.**
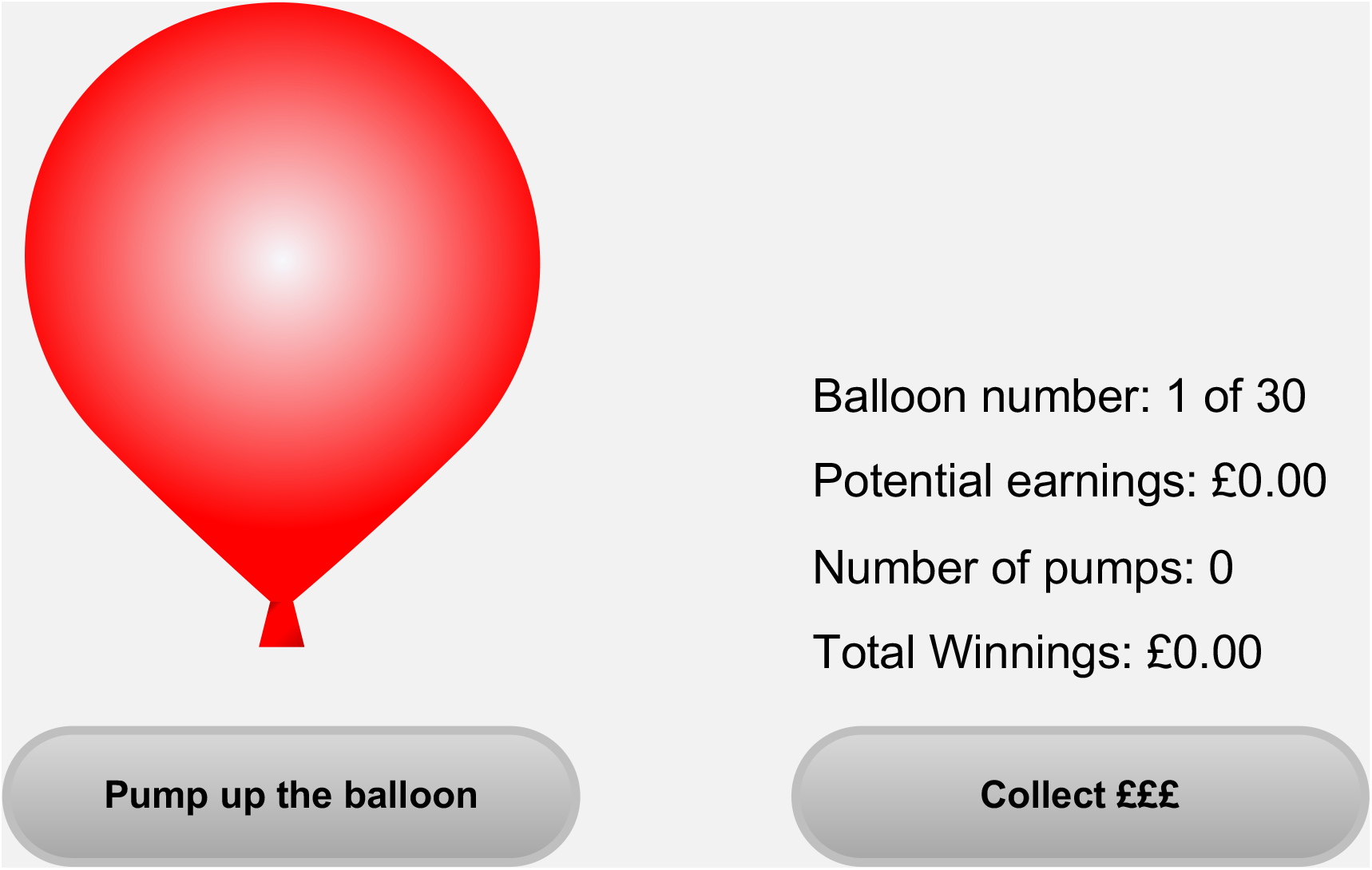
Visual display of the Balloon Analogue Risk Task. ‘Pump up the balloon’ and ‘Collect £££’ are the two available response options. Visual feedback of ‘Balloon number’, ‘Potential earnings’, ‘Number of pumps’ and ‘Total winnings’ are displayed throughout each trial.

## Cognitive Function

### Central Nervous System Vital Signs (CNSVS)

CNSVS is a computerised neurocognitive test battery comprising of neuropsychological tests to assess cognitive behaviour and acts as a tool, not for diagnosis, but for brief clinical evaluation of mild cognitive dysfunction (Gualtieri & Johnson, 2006). Participants completed all tests on their computer and made their responses using a keyboard. The scores produced from these tests contribute to neurocognitive clinical evaluation domains. Nine tests were included within the current research which were linked to 14 cognitive domains: composite memory, verbal memory, visual memory, psychomotor speed, reaction time, complex attention, cognitive flexibility, processing speed, executive function, reasoning, working memory, sustained attention, simple attention and motor speed. Automated scoring reported raw patient test scores for each domain which were automatically normalised and age-matched to a large normative database to create standard scores. These scores were produced for the 14 domains along with the neurocognitive index (NCI) which represents a global score of neurocognition by taking an average of the domain scores for composite memory, psychomotor speed, reaction time, complex attention and cognitive flexibility. Standard scores for NCI and working memory were included in analyses.

### Genetic Data

Five specific genetic polymorphisms which formed the DGRS were identified for each participant. Genetic analysis was conducted by LGC Genomics, and full methodology can be found at: http://www.lgcgenomics.com/. The single nucleotide polymorphisms within four genes were determined using kompetitive allele specific polymerase change reaction (KASP PCR) genotyping: DRD1 (rs4532), DRD2 (rs1800497), DRD3 (rs6280) and COMT (rs4680). This process produced a bi-allelic score for each single nucleotide polymorphism. The variable number tandem repeat in the DAT gene (rs28363170) was analyzed using a separate PCR process. Here, the PCR was followed by PCR clean-up, sanger sequencing and genotype calling. The repeat length of DAT VNTR was determined by eye on the sequence trace files.

Dependent upon the specific mutation/number of repeats for each of the five polymorphisms, every participant received a score of 0 or 1 for each polymorphism according to whether it acts to decrease or increase dopamine transmission, respectively (Pearson-Fuhrhop et al. 2013, 2014, MacDonald et al. 2016). All gene scores were then summed for an overall DGRS between 0-5 (higher score = higher dopamine levels) (Table 1S, Online Resource). All genes were in Hardy-Weinberg equilibrium (all p > .291), which was determined with chi-square tests. For the linear regression models discussed below, the sample size for each DGRS was as follows: DGRS 0 n = 0; DGRS 1 n = 4, DGRS 2 n = 11, DGRS 3 n = 17, DGRS 4 n = 4, DGRS 5 n = 14. The DGRS was split into two groups: DGRS low (DGRS 0-2) and DGRS high (DGRS 3-5) aiming to make as equal sample sizes as possible. The DGRS was utilised as a binary independent variable within the models.

**Table 1.** Participant clinical, demographic, genetic and cognitive variables separated by incidence of impulse control behaviours via the QUIP-short.

| **Dopamine Agonist (DA)** |  |  |  |
| --- | --- | --- | --- |
|  | **ICB (n = 28)** | **No ICB (n = 30)** | **p** |
| Age, years | 63.3 (8.83) | 64.7 (8.87) | .546 |
| **BIS percentage** | **52.5 (10.5)** | **44.8 (7.85)** | **.002** |
| CNSVS NCI | 87.9 (28.0) | 97.7 (10.3) | .092 |
| CNSVS WM | 98.3 (20.8) | 102 (17.8) | .436 |
| DA LEDD | 210 (110) | 190 (119) | .495 |
| DA type, % ropinirole<br>(n, ropinirole:pramipexole:rotigotine) | 57.1 (16:8:4) | 70.0 (21:5:4) | .477 |
| **DGRS** | 3.21 (1.17) | 3.07 (1.01) | .608 |
| **Gender, % male (n, male:female)** | **67.9 (19:9)** | **33.3 (10:20)** | **.008** |
| **ICB frequency (QUIP-RS)** | **26.3 (12.6)** | **7.90 (9.94)** | **.010 ♦** |
| **ICB frequency (QUIP-short)** | **2.50 (1.32)** | **0** | **<.001 ♦** |
| Total LEDD | 684 (431) | 677 (596) | .961 |
| UPDRS I&II | 22.6 (11.6) | 17 (10.3) | .057<sup>^</sup> |
| Years on DA | 5.57 (4.01) | 4.58 (3.31) | .309 |
| Years since diagnosis | 8.07 (5.79) | 6.97 (4.67) | .426 |
| **Non-Dopamine Agonist (NDA)** |  |  |  |
|  | **ICB (n = 11)** | **No ICB (n = 14)** | **p** |
| Age, years | 62.6 (9.67) | 66.1 (7.68) | .332 |
| BIS percentage | 52.2 (9.44) | 46.6 (8.30) | .133 |
| CNSVS NCI | 84.9 (20.4) | 93.5 (16.4) | .251 |
| CNSVS WM | 100.8 (12.4) | 99.9 (14.3) | .861 |
| DGRS | 3.09 (1.14) | 3.43 (1.16) | .473 |
| Gender, % male (n, male:female) | 81.8 (9:2) | 64.3 (9:5) | .353 |
| **ICB Score (QUIP RS)** | **24.8 (17.5)** | **9.43 (8.53)** | **.008** |
| **ICB Score (QUIP-short)** | **2.64 (1.75)** | **0** | **<.001 ♦** |
| Total LEDD | 665 (520) | 350 (271) | .062<sup>^</sup> |
| UPDRS I&II | 20.9 (10.6) | 14.9 (9.08) | .372 |
| **Years since diagnosis** | **4.64 (2.73)** | **2.68 (1.73)** | **.039** |

Means for variables ( ± standard deviation). ICB: impulse control behaviour (n: number); BIS percentage: Barratt impulsiveness scale; CNSVS: central nervous system vital signs; NCI: neurocognitive index; WM: working memory; DA: dopamine agonist; LEDD: levodopa equivalent daily dose; DGRS: Dopamine Genetic Risk Score; QUIP: Questionnaire for impulsive-Compulsive Disorders in Parkinson's Disease; RS: rating scale; UPDRS: Unified Parkinson's Disease Rating Scale; Significant values in bold ( $p < .05$ ). ♦: Wilcoxon rank sum test. ^: trending towards significance.

### Statistical Analysis

Statistical analysis and modelling were performed in MATLAB (version R2020a, MathWorks). As a preliminary analysis, comparisons were made for all available clinical, demographic, genetic and cognitive variables between those with and without an ICB. Seventeen participants with unavailable data for these variables due to errors in reporting and incomplete online datasets from the CNSVS were discarded from these analyses (DA n = 12, NDA n = 5). Kolmogorov-Smirnov tests identified any violations of normality. Wilcoxon rank sum tests were used to compare any variables which violated normality, while the remaining variables were compared using unpaired t-tests. A simple linear regression looked for a correlation between ICB frequency on the QUIP-RS and BIS score in both DA and NDA groups. The following linear regression models identified the variables associated with clinical and trait impulsivity.

### Clinical Impulsivity model

The response variable for this model was ICBs identified via the QUIP. A participant’s score on the QUIP-S and QUIP-RS were strongly correlated (R = 0.72, p < .001). Therefore, we chose to predict results of the QUIP-RS because a larger scale range was likely to be more sensitive to changes in impulsivity. CNSVS NCI and WM were not included due to missing data, as their inclusion would have reduced the sample size of the model. DGRS, DA levodopa equivalent daily dose (DA LEDD), Negative Reinforcement from the BART, SSRT from Stop Both trials of the ARIT, and Years on DA were selected a-priori to be included in the model to test our hypotheses and build on previous literature (MacDonald et al. 2016; Hall et al. 2021). Univariate linear regression analyses identified any additional variables which could be included as independent predictors of ICB frequency in the full model (Table 2S, Online Resource). However, any continuous variables identified were tested for collinearity against the pre-selected variables, and resultant correlated variables were not included in the final model (Table 3S, Online Resource). Therefore, UPDRS I&II and Years Since Diagnosis were not included in the final model as they both correlated with Years on DA (both p < 0.001). Gender was also not included to not overparameterise the model. The final mixed-effects multiple linear regression model was formed with selected variables and hypothesised interactions:

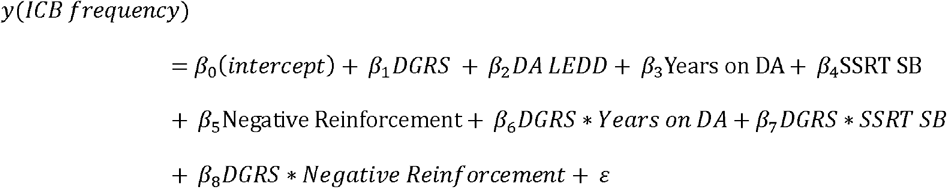

**Table 2.** Multiple linear regression analysis of variables associated with the frequency of impulse control behaviours.

| **ICB (n = 23) no ICB (n = 27)** |  |  |  |  |
| --- | --- | --- | --- | --- |
|  | **β** | **SE** | **p value** | **95 % CI (β)** |
| Intercept | -15.0 | 13.0 | .254 | [-41.3, 11.2] |
| DGRS low | 9.09 | 24.1 | .708 | [-39.5, 57.7] |
| LEDD DA | -0.004 | 0.02 | .862 | [-0.04, 0.04] |
| **Negative Reinforcement** | **12.3** | **4.76** | **.014** | **[2.63, 21.9]** |
| SSRT stop both | 0.07 | 0.03 | .056^ | [-0.002, 0.14] |
| **Years on DA** | **2.09** | **0.48** | **<.001** | **[1.11, 3.06]** |
| DGRS low * Negative Reinforcement | 11.5 | 15.5 | .463 | [-19.8, 42.9] |
| DGRS low * SSRT stop both | 0.009 | 0.08 | .911 | [-0.15, 0.17] |
| DGRS low * Years on DA | -1.19 | 1.15 | .305 | [-3.52, 1.13] |

Response variable: score on Questionnaire for Impulsive-Compulsive Disorders in Parkinson's Disease rating scale. ICB: impulse control behaviour (n: number); DGRS: dopamine genetic risk score; LEDD: levodopa equivalent daily dose; DA: Dopamine Agonist; SSRT: stop signal reaction time; β: coefficient, SE: standard error, CI: confidence interval. Significant values in bold ( $p < .05$ ). ^: trending towards significance.

**Table 3.** Multiple linear regression analysis of variables associated with Barratt Response variable: Barratt Impulsiveness Scale percentage.

| **ICB (n = 23) no ICB (n = 27)** |  |  |  |  |
| --- | --- | --- | --- | --- |
|  | **$\beta$** | **SE** | **p value** | **95 % CI ($\beta$)** |
| **Intercept** | **38.8** | **9.56** | **<.001** | **[19.5, 58.1]** |
| DGRS low | -4.37 | 17.7 | .806 | [-40.1, 31.4] |
| LEDD DA | -0.008 | 0.02 | .582 | [-0.04, 0.02] |
| Negative Reinforcement | 1.79 | 3.50 | .612 | [-5.28, 8.87] |
| SSRT stop both | 0.02 | 0.03 | .358 | [-0.03, 0.08] |
| **Years on DA** | **1.14** | **0.36** | **.003** | **[0.42, 1.86]** |
| DGRS low * Negative Reinforcement | -1.19 | 11.4 | .917 | [-24.2, 21.9] |
| DGRS low * SSRT stop both | 0.004 | 0.06 | .948 | [-0.12, 0.12] |
| DGRS low * Years on DA | 0.24 | 0.85 | .780 | [-1.47, 1.95] |

Response variable: Barratt Impulsiveness Scale percentage. ICB: impulse control behaviour (n: number); DGRS: dopamine genetic risk score; LEDD: levodopa equivalent daily dose; DA: Dopamine Agonist; SSRT: stop signal reaction time; $\beta$ : coefficient, SE: standard error, CI: confidence interval. Significant values in bold ( $p < .05$ ).

Further linear regressions were run with this model to determine the contribution of each individual genetic polymorphism towards the response variable. This involved substituting the score (0 or 1) for each genetic polymorphism into the model in place of the full DGRS. The same model was run for the NDA group, without DA LEDD and Years on DA.

### Trait Impulsivity model

The same independent variables and interactions from the clinical impulsivity model were selected for inclusion in the multiple linear regression model predicting BIS percentage as the response variable:

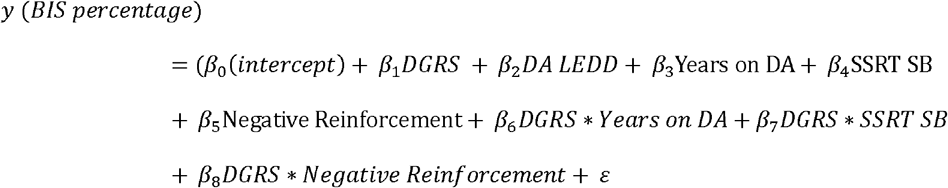

The same model was run for the NDA group, without DA LEDD and Years on DA.

### Model Validation

Effect sizes for all models were determined and interpreted using adjusted R^2^ (0.01 = small, 0.09 = medium, 0.25 = large, Foster et al., 2018) and the achieved statistical power is reported (G*Power 3.1.9.6). Validation against a constant model (i.e., goodness-of-fit) was assessed for all models and an alpha value of 0.05 was used for all analyses.

## RESULTS

### Preliminary Analysis

Data from 83 participants (DA: n = 58, 45-77 years, mean 64.1 ± 8.80 standard deviation, NDA: n = 25, 46-79 years, mean 64.6 ± 8.60 standard deviation) were included in the preliminary clinical, demographic, genetic and cognitive comparisons between those with (QUIP-S > 1) and without (QUIP-S = 0) an ICB (Table 1). Of these participants, in the DA group, 16 participants had a low DGRS (0-2) and 42 had a high DGRS (3-5). Moreover, in the NDA group, 9 participants had a low DGRS and the remaining 16 presented a high DGRS. In the DA group, participants with an ICB were more likely to be male (p = .008) and presented with a higher BIS (p = .002) and QUIP-RS (p = .010) score. Scores on the UPDRS I&II trended towards being higher for those with an ICB than those without. These results were not likely to be due to changes in general cognitive function as there were no differences between CNSVS NCI and WM between ICB groups. In the NDA group, those with an ICB reported a greater number of years since diagnosis (p = .039),a higher QUIP-RS (p = .008) score, and the increased overall medication dosage (Total LEDD) trended towards significance (p = .062).

### Linear Regression Models

The sample sizes for the following models were reduced (DA n = 50, NDA n = 22) due to incomplete datasets for included independent variables or the inability to genotype from the DNA sample. The following results are specific to DA medication, as NDA models were unable to explain any variability in the outcome variable (goodness-of-fit: clinical model p = .951, trait model p = .662). There was therefore nothing to report for these NDA models.

### Clinical Impulsivity

#### Task performance and exposure time to DA medication were associated with the frequency of ICBs

The Clinical Impulsivity model (Table 2) was validated against a constant model (F_7,41_ = 3.15, p = .007) and explained 26% of the variance in ICB frequency scores according to the adjusted R^2^ value (unadjusted R^2^ = 0.381, i.e., large effect size). The statistical power achieved by the model was 97.2%, also indicating an appropriate sample size for the model. ICB frequency increased by 12.3 for every 1 unit increase in negative reinforcement (b = 12.4, p = .014). This statistic indicates that, as expected, people who made more impulsive decisions on the BART after a loss also reported a higher frequency of ICBs. The increase in ICB frequency of 0.07 for each millisecond increase in SSRT SB trended towards significance (b = 0.07, 0 = .056), indicating people with worse motor impulsivity tended to report a higher frequency of ICBs, as predicted. As hypothesised, ICB frequency increased by 2.09 for every year on DA medication (b = 2.09, p <.001). However, contrary to our hypotheses, these associations between clinical impulsivity and task performance/time on medication did not depend on a participant’s DGRS (DGRS X Negative Reinforcement: b = 11.5, p = 0.463; DGRS X SSRT: b = 0.009, p = 0.911; DGRS X Years on DA: b = -1.19, p = 0.305). DA dose (b = -0.004, p = 0.862) and DGRS alone (b = 9.09, p = 0.708) were also not predictive of ICB frequency score.

Interestingly, two of the DGRS constituent genes interacted with time on DAs to effect ICB frequency. When substituting COMT into the model (F_7,41_ = 4.1, p = .001, R^2^ = 0.444 i.e., large effect size, 95.7% power), the increase in ICB frequency from one year on DAs was 2.49 more for a COMT score of 1 (greater dopamine neurotransmission, b = 2.12) compared to 0 (b = -0.37, p = .048). Similarly for DAT (F_7,41_ = 4.57, p <.001, R^2^ = 0.471 i.e., large effect size, 95.6% power), the increase in ICB frequency from one year on DAs was 1.14 more for a DAT score of 1 (b = 2.56) compared to 0 (b =1.42, p = .014). DAT score also interacted with Negative Reinforcement. For participants with a DAT score of 1, a single unit increase in Negative Reinforcement reduced ICB frequency by 28 (b = -11.0) compared to participants with a score of 0 (b = 17.0, p = 026). No individual genetic polymorphism was independently associated with a change in ICB frequency (p > .288).

### Trait Impulsivity

Trait impulsivity (BIS percentage) was significantly correlated with clinical impulsivity (ICB frequency) in both DA (R = 0.56, p <.001, Figure 3) and NDA (R = 0.74, p<.001, Figure 4) groups. This indicates that participants who reported higher levels of everyday trait impulsivity, also reported a higher frequency of ICBs.

**Fig. 3.**
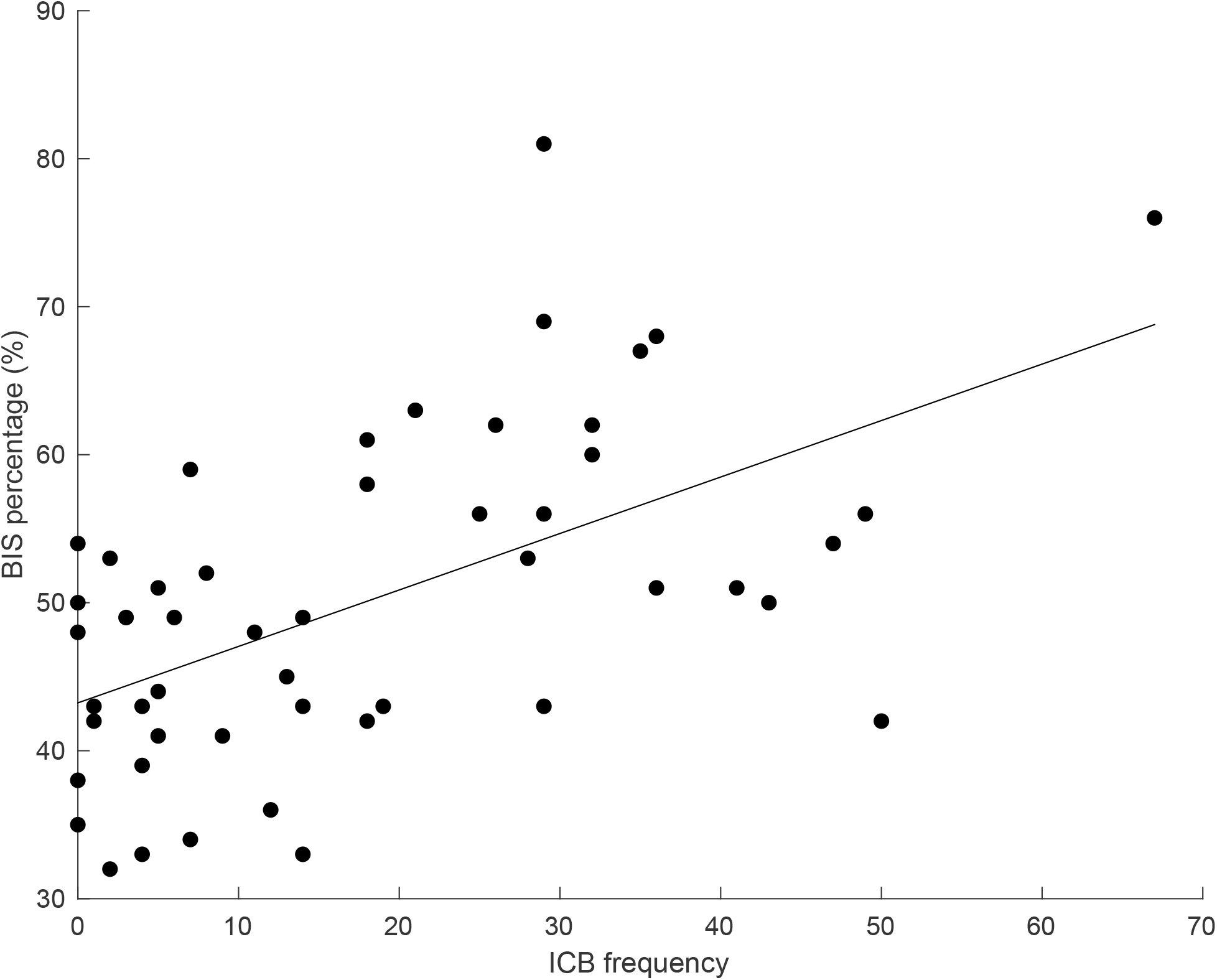
Linear correlation between impulse control behaviour (ICB) frequency measured via the QUIP-RS and Barratt Impulsiveness Scale (BIS) percentage score in the dopamine agonist group. Data circles represent individual participants.

**Fig. 4.**
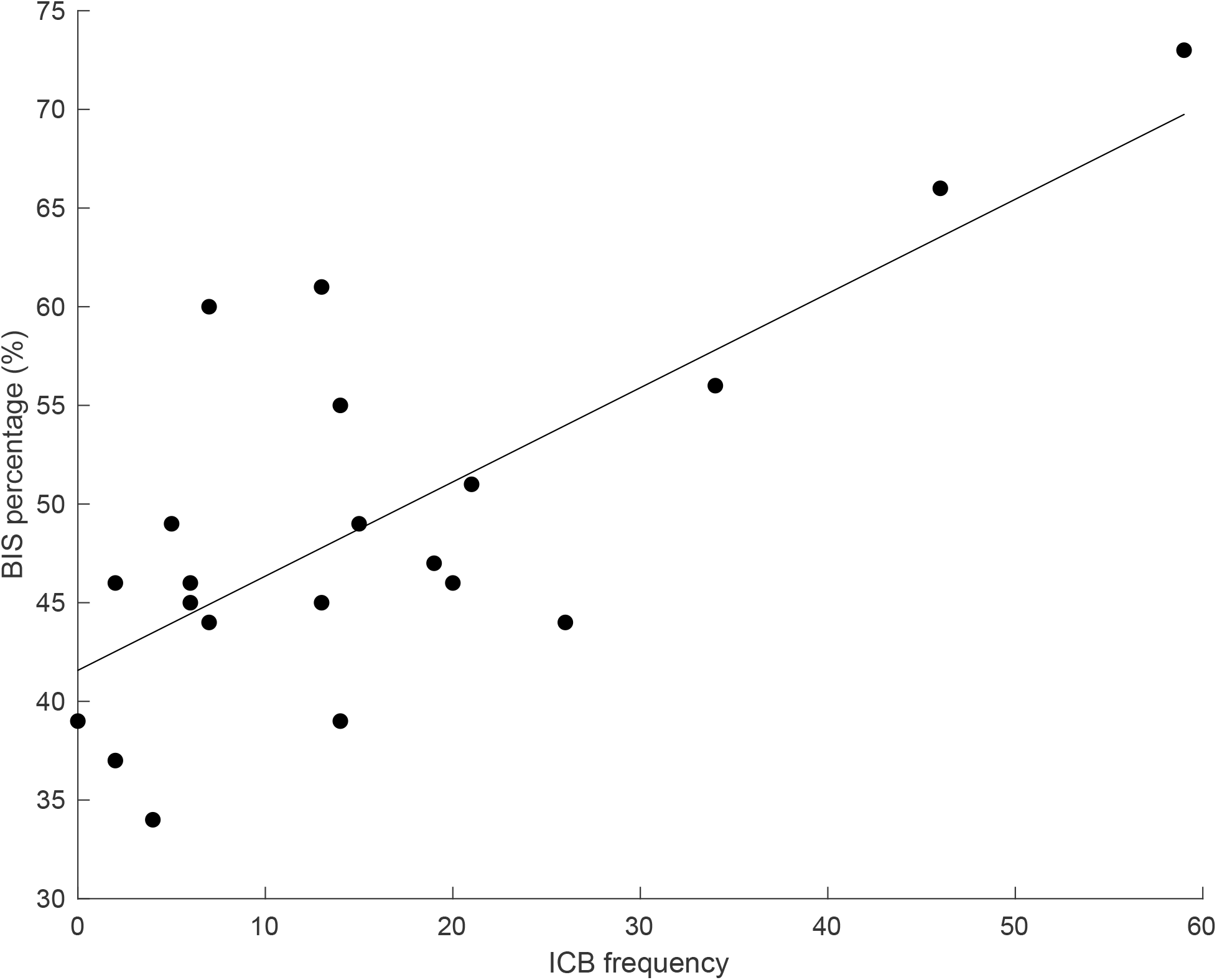
Linear correlation between impulse control behaviour (ICB) frequency measured via the QUIP-RS and Barratt Impulsiveness Scale (BIS) percentage score in the non-dopamine agonist group. Data circles represent individual participants.

### Long term exposure to DA medication predicted subjective, real-world trait impulsivity

For trait impulsivity (F_7,41_ = 1.98, p = .074, R^2^ = 0.28 i.e., large effect size, 83.5% power, Table 3), a participant’s BIS increased by 1.14% with every year on DA medication (b = 1.14, p = .003). No other independent variables or interactions significantly predicted BIS percentage (p > .358).

## DISCUSSION

The primary focus of this study was to investigate the sensitivity of objective task measures, along with variation in dopamine genetics, to determine the frequency of clinically identified ICBs. As such, the study produced several novel findings which were specific to DA medication. As hypothesised, task performance was associated with ICBs. Participants who made a greater number of impulsive decisions after a loss on the BART, or who tended to exhibit worse impulsivity on the ARIT, also reported a higher frequency of impulsive behaviours on the clinical screening tool. However, contrary to the other aspect of our hypotheses, DGRS, an analogy of dopamine neurotransmission, did not interact with task performance to determine clinical impulsivity. Interestingly, the DAT polymorphism interacted with impulsive decision making on the BART to effect ICB frequency. The secondary aim of this study was to work towards identifying measures for prognostic use for ICBs on dopamine agonists, thus time on DA medication and DA dosage were incorporated into the models. Greater length of exposure to DA medication was associated with higher ICB frequency as predicted, whereas DA dosage was not. The DGRS did not interact with time on DAs, however when examining the influence of individual genes, more dopamine neurotransmission indexed via polymorphisms in COMT and DAT predicted higher ICB frequency with increasing exposure to DA medication. More time on DA medication was also associated with higher levels of trait impulsivity, which in turn was correlated with ICB frequency. The results of the current study present promising initial results highlighting the potential use of our task-derived measures of impulse control to predict ICB severity in people with Parkinson’s disease on DA medication.

A linear relationship existed between task performance and clinically identified ICBs, but only for patients taking dopamine agonist medication. Patients who made more impulsive decisions on the BART after a loss also reported a higher frequency of ICBs. Our finding aligns with other studies that show ICB patients failing to reduce their impulsive behaviour following a loss on the BART, reflecting punishment (Martini et al. 2018), although this effect has not been previously confirmed to be agonist specific. However, when negative feedback is calculated slightly differently as the difference between number of balloon pumps directly preceding and following a loss, PD patients can show reduced impulsive behaviour irrespective of ICB and DA status (Claassen et al. 2011). For performance on the ARIT in our study, worse impulsive action (a longer SSRT) tended to be associated with increased ICB frequency. To our knowledge, we are the first to investigate the relationship between ICBs and ARIT performance. Findings using SSRT derived from the stop signal task have been mixed. Studies have reported no differences in SSRT between PD patients with and without ICBs (Ricciardi et al. 2017; Vriend et al. 2018; Hlavata et al. 2020), as well as shorter SSRTs in ICB patients compared not only to PD patients without ICBs, but also to healthy control participants (Claassen et al. 2015). The positive relationship between SSRT and ICB frequency in our study may be due to task design, as the ARIT explores control of internally generated, rather than the externally cued responses. PD patients find internally generated responses with an anticipatory component most difficult (Jahanshahi et al. 1995), which likely reflects a sensitivity of predictive timing processes to the ongoing deterioration of the prefrontal-basal ganglia network (Cunnington et al. 1995) and therefore potentially dopaminergic MCL function. Overall, our objective task measures show promise as sensitive markers of impulsivity problems on DAs leading to real-world impulsive behaviours. It remains to be seen whether impaired task performance precedes ICB development.

Contrary to our hypotheses, there was no association between DGRS and ICB frequency and no interaction between task measures and the full DGRS. This finding contrasts with our previous finding that the DGRS can explain the incidence of ICBs (Hall et al. 2021). However, there is a key distinction between the studies. Namely, our previous study was predicting the binary presence/absence of any ICB, whereas the current study tried to link the DGRS with a measure closer to ICB severity i.e., frequency of ICBs. The rationale for this was twofold: 1) to use the more finely grained and wider ranging responses on the QUIP-RS (compared to the QUIP-S) for maximal sensitivity to subtle changes in task measures e.g., a change in SSRT of a few milliseconds, and 2) because the smaller sample size in a binary outcome variable in the current study would have limited overall model sensitivity (23 QUIP-S ≥ 1, 27 = 0). Combined, perhaps our results speak to the DGRS being able to predict the development of an ICB, rather than determining the more subtle distinction between severity of behaviours. Interestingly, although the full DGRS did not interact with task measurers, the DAT polymorphism in isolation interacted with impulsive decision making on the BART to effect ICB frequency. A relationship between DAT and cognitive impulsivity task performance has previously been reported (Mata et al. 2012; MacDonald et al. 2016). DAT is responsible for the reuptake of dopamine into pre-synaptic neurons (Hovde et al. 2019) and predominantly removes dopamine from within the striatum, a key region for cognitive decision making (Mata et al. 2012; Vriend et al. 2014). A higher DAT score represents a less functional DAT protein, which leads to less clearance of dopamine from the synaptic cleft, and greater striatal dopamine neurotransmission (Cilia et al. 2010; Vriend et al. 2014). In our study, patients with higher striatal dopamine levels (i.e., DAT = 1) who made more impulsive decisions on the BART counterintuitively had lower, rather than higher, clinical impulsivity. There is no immediately obvious reason for this paradoxical finding, but it should be interpreted with caution, as the study was not designed to primarily investigate single gene effects.

Increased exposure to DA medication, but not increasing dose, predicted higher trait impulsivity and increased ICB frequency. The effect of purely time on DAs separate from dose has not been widely reported. Of those who did isolate time on DAs, some studies reported a positive correlation with ICBs (Giladi et al. 2007; Corvol et al. 2018), whereas others did not (Bastiaens et al. 2013). The findings for DA dose are also somewhat mixed, although a greater proportion of studies have previously determined a positive association between DA dosage and ICBs (Weintraub et al. 2006; Lee et al. 2010; Joutsa et al. 2012; Perez-Lloret et al. 2012; Bastiaens et al. 2013; Corvol et al. 2018; Markovic et al. 2020), than no relationship (Isaias et al. 2008; Housden et al. 2010; Weintraub et al. 2010; Callesen et al. 2014; Vela et al. 2016). The reduced D2 auto-receptor sensitivity hypothesis explained previously is one potential neural mechanisms of action underlying our effect of time on DAs. Epigenetics may also be playing a role. Dopamine medication may regulate DNA transcription over time to increase protein and therefore neurotransmitter production (Lepack et al. 2020), potentially leading to the increase in impulsive behaviour. In our study, COMT and DAT mutations resulting in greater dopamine neurotransmission were associated with higher ICB frequency with increasing time on DA medication. Again single-gene exploratory findings should be interpreted with caution but could point to future epigenetics work including these genes when investigating gene vs medication interactions in the context of ICB severity over time.

Participants who reported higher levels of everyday trait impulsivity, also reported a higher frequency of ICBs in both the DA and NDA groups. Impulsive trait behaviour is a risk factor for ICBs (Leeman & Potenza, 2011; Weintraub & Mamikonyan, 2019) and PD patients with ICBs have reported higher impulsivity on the BIS compared to those without ICBs (Isaias et al. 2008; Marin-Lahoz et al. 2018; Hlavata et al. 2020; Takahashi et al. 2022). Our positive correlation between BIS and QUIP-RS in both DA and NDA groups has previously been reported in a group of PD patients, but it is uncertain how many of these patients were on DA medication (Goerlich-Dobre et al. 2014). The presence of a comparable relationship in both groups suggests that the behavioural manifestation of ICBs in an NDA group may be similar to those on DAs. However, our clinical model was unable to account for the variability in ICB severity for this group, indicating the underlying mechanisms for ICBs may be distinct for agonist vs non-agonist medication (Kelly et al. 2020).

It is important to acknowledge some limitations of the current study. Firstly, the NDA control group had a smaller sample size than the DA group due to recruitment time constraints. The smaller sample size and reduced variability may have contributed to our clinical model being unable to account for ICB frequency in the NDA group. Although it is worth noting the ICB variability was still sufficient to reveal a correlation with BIS scores, and the NDA group reported a similar average and range of QUIP scores compared to the DA group. Nevertheless, future work should aim to replicate this lack of effect with the clinical model in a larger group of PD patients who are taking only non-agonist medication. Secondly, although we present novel findings by including time on DA medication in our models, this was a cross sectional study. A longitudinal study design is required to confirm interactions with time on an individual basis. A longitudinal design would also reveal whether task performance tracks with ICB changes over time. If this design was conducted with de novo patients, it could additionally reveal any changes to predictive variables that precede increases in ICBs, which is a crucial step towards identifying measures for prognostic use.

In summary, this study provides evidence that objective measures from impulse control tasks and time of exposure to medication can explain ICB severity in people with PD and are specific to DA mechanisms of effect. On the other hand, the DGRS appears better suited to predicting the incidence, rather than severity, of ICBs on DAs. It remains to be determined whether task performance can be used to monitor ICB changes over time within an individual on agonist medication.

## Data Availability

All data produced in the present study are available upon reasonable request to the authors

## STATEMENTS AND DECLARATIONS

### Ethical approval and informed written consent

Ethical approval was acquired from The University of Birmingham Ethics Committee to complete the current study, and informed consent was obtained from all individual participants included in the study.

### Financial Disclosure/Conflict of Interest concerning the research related to the manuscript

The authors declare no conflicts of interest.

### Author’s Roles

AH and HM participated in all aspects of the development of the research project, data/statistical analyses and the writing and revisions of the manuscript. MW participated in data/statistical analyses and the review and critique of the final manuscript. NJ participated in the development of the research project, and the review and critique of the final manuscript.

## REFERENCES

Abidin, S. Z., Tan, E. L., Chan, S. C., Jaafar, A., Lee, A. X., Abd Hamid, M. H. N., Abdul Murad, N. A., Pakarul Razy, N. F., Azmin, S., Ahmad Annuar, A., Lim, S. Y., Cheah, P. S., Ling, K. H., & Mohamed Ibrahim, N. (2015). DRD and GRIN2B polymorphisms and their association with the development of impulse control behaviour among Malaysian Parkinson’s disease patients. BMC Neurology, 15(1), 1–10. https://doi.org/10.1186/s12883-015-0316-2

Bastiaens, J., Dorfman, B. J., Christos, P. J., & Nirenberg, M. J. (2013). Prospective Cohort Study of Impulse Control Disorders in Parkinson’s Disease. Movement Disorders, 28(3), 327–333. https://doi.org/10.1002/mds.25291

Callesen, M. B., Weintraub, D., Damholdt, M. F., & Møller, A. (2014). Impulsive and compulsive behaviors among Danish patients with Parkinson’s disease: Prevalence, depression, and personality. Parkinsonism and Related Disorders, 20(1), 22–26. https://doi.org/10.1016/j.parkreldis.2013.09.006

Caminiti, S. P., Presotto, L., Baroncini, D., Garibotto, V., Moresco, R. M., Gianolli, L., Volonté, M. A., Antonini, A., & Perani, D. (2017). Axonal damage and loss of connectivity in nigrostriatal and mesolimbic dopamine pathways in early Parkinson’s disease. NeuroImage: Clinical, 14(October 2016), 734–740. https://doi.org/10.1016/j.nicl.2017.03.011

Cilia, R., Ko, J. H., Cho, S. S., van Eimeren, T., Marotta, G., Pellecchia, G., Pezzoli, G., Antonini, A., & Strafella, A. P. (2010). Reduced dopamine transporter density in the ventral striatum of patients with Parkinson’s disease and pathological gambling. Neurobiology of Disease, 39(1), 98–104. https://doi.org/10.1016/j.nbd.2010.03.013

Claassen, D. O., Wildenberg, W. P. M. van den, Ridderinkhof, K. R., Jessup, C. K., Harrison, M. B., Wooten, G. F., & Scott A. Wylie. (2011). The Risky Business of Dopamine Agonists in Parkinson Disease and Impulse Control Disorders. Behavioural Neuroscience, 125(4), 492–500. https://doi.org/10.1037/a0023795.

The Claassen, D. O., Van Den Wildenberg, W. P. M., Harrison, M. B., Van Wouwe, N. C., Kanoff, K., Neimat, J. S., & Wylie, S. A. (2015). Proficient motor impulse control in Parkinson disease patients with impulsive and compulsive behaviors. Pharmacology Biochemistry and Behavior, 129, 19–25. https://doi.org/10.1016/j.pbb.2014.11.017

Claassen, D. O., Stark, A. J., Spears, C. A., Petersen, K. J., van Wouwe, N. C., Kessler, R. M., Zald, D. H., & Donahue, M. J. (2017). Mesocorticolimbic hemodynamic response in Parkinson’s disease patients with compulsive behaviors. Movement Disorders, 32(11), 1574–1583. https://doi.org/10.1002/mds.27047

Congdon, E., Constable, R. T., Lesch, K. P., & Canli, T. (2009). Influence of SLC6A3 and COMT variation on neural activation during response inhibition. Biological Psychology, 81(3), 144–152. https://doi.org/10.1016/j.biopsycho.2009.03.005

Cools, R., Barker, R. A., Sahakian, B. J., & Robbins, T. W. (2001). Enhanced or impaired cognitive function in Parkinson’s disease as a function of dopaminergic medication and task demands. Cerebral Cortex, 11(12), 1136–1143. https://doi.org/10.1093/cercor/11.12.1136

Cools, R. (2006). Dopaminergic modulation of cognitive function-implications for L-DOPA treatment in Parkinson’s disease. In Neuroscience and Biobehavioral Reviews (vol. 30, Issue 1, pp. 1–23). https://doi.org/10.1016/j.neubiorev.2005.03.024

Corvol, J. C., Artaud, F., Cormier-Dequaire, F., Rascol, O., Durif, F., Derkinderen, P., Marques, A. R., Bourdain, F., Brandel, J. P., Pico, F., Lacomblez, L., Bonnet, C., Brefel-Courbon, C., Ory-Magne, F., Grabli, D., Klebe, S., Mangone, G., You, H., Mesnage, V., … Elbaz, A. (2018). Longitudinal analysis of impulse control disorders in Parkinson disease. Neurology, 91(3), e189–e201. https://doi.org/10.1212/WNL.0000000000005816

Cunnington R, Iansek R, Bradshaw JL, Phillips JG. (1995) Movement- related potentials in Parkinson’s disease. Presence and predictability of temporal and spatial cues. Brain, 118(4), 935–950. https://doi.org/10.1093/brain/118.4.935

Dauer, W., & Przedborski, S. (2003). Models and mechanisms. Neuron, 39, 889–909. https://doi.org/10.1016/s0896-6273(03)00568-3

Erga, A. H., Dalen, I., Ushakova, A., Chung, J., Tzoulis, C., Tysnes, O. B., Alves, G., Pedersen, K. F., & Maple-Grødem, J. (2018). Dopaminergic and Opioid Pathways associated with impulse control Disorders in Parkinson’s Disease. Frontiers in Neurology, 9(109). https://doi.org/10.3389/fneur.2018.00109

Faouzi, J., Couvy-Duchesne, B., Bekadar, S., Colliot, O., & Corvol, J. C. (2021). Exploratory analysis of the genetics of impulse control disorders in Parkinson’s disease using genetic risk scores. Parkinsonism and Related Disorders, 86, 74–77. https://doi.org/10.1016/j.parkreldis.2021.04.003

Ford, C. P. (2014). The role of D2-autoreceptors in regulating dopamine neuron activity and transmission. Neuroscience, 282, 13–22. https://doi.org/10.1016/j.neuroscience.2014.01.025

Gasser, T., Wichmann, T., & DeLong, M. R. (2015). Parkinson Disease and Other Synucleinopathies. In Neurobiology of Brain Disorders: Biological Basis of Neurological and Psychiatric Disorders (pp. 281–302). https://doi.org/10.1016/B978-0-12-398270-4.00019-7

Gatto, E. M., & Aldinio, V. (2019). Impulse Control Disorders in Parkinson’s Disease. A Brief and Comprehensive Review. Frontiers in Neurology, 10(351). https://doi.org/10.3389/fneur.2019.00351

Giladi, N., Weitzman, N., Schreiber, S., Shabtai, H., & Peretz, C. (2007). New onset heightened interest or drive for gambling, shopping, eating or sexual activity in patients with Parkinson’s disease: The role of dopamine agonist treatment and age at motor symptoms onset. Journal of Psychopharmacology, 21(5), 501–506. https://doi.org/10.1177/0269881106073109

Goerlich-Dobre, K. S., Probst, C., Winter, L., Witt, K., Deuschl, G., Möller, B., & Van Eimeren, T. (2014). Alexithymia-an independent risk factor for impulsive-compulsive disorders in Parkinson’s disease. Movement Disorders, 29(2), 214–220. https://doi.org/10.1002/mds.25679

Gualtieri, C. T., & Johnson, L. G. (2006). Reliability and validity of a computerized neurocognitive test battery, CNS Vital Signs. Archives of Clinical Neuropsychology, 21(7), 623–643. https://doi.org/10.1016/j.acn.2006.05.007

Hall, A., Weaver, S. R., Compton, L. J., Byblow, W. D., Jenkinson, N., & MacDonald, H. J. (2021). Dopamine genetic risk score predicts impulse control behaviors in Parkinson’s disease. Clinical Parkinsonism & Related Disorders, 5(August), 100113. https://doi.org/10.1016/j.prdoa.2021.100113

Hlavatá, P., Linhartová, P., Šumec, R., Filip, P., Světlák, M., Baláž, M., Kašpárek, T., & Bareš, M. (2020). Behavioral and Neuroanatomical Account of Impulsivity in Parkinson’s Disease. Frontiers in Neurology, 10. https://doi.org/10.3389/fneur.2019.01338

Housden, C. R., O’Sullivan, S. S., Joyce, E. M., Lees, A. J., & Roiser, J. P. (2010). Intact reward learning but elevated delay discounting in Parkinson’s disease patients with impulsive-compulsive spectrum behaviors. Neuropsychopharmacology, 35(11), 2155–2164. https://doi.org/10.1038/npp.2010.84

Hovde, M. J., Larson, G. H., Vaughan, R. A., & Foster, J. D. (2019). Model systems for analysis of dopamine transporter function and regulation. Neurochemistry International, 123(August 2018), 13–21. https://doi.org/10.1016/j.neuint.2018.08.015

Ihle, J., Artaud, F., Bekadar, S., Mangone, G., Sambin, S., Mariani, L. L., Bertrand, H., Rascol, O., Durif, F., Derkinderen, P., Scherzer, C., Elbaz, A., Corvol, J. C., Corvol, J. C., Elbaz, A., Vidailhet, M., Brice, A., Artaud, F., Bourdain, F., … Faurie-Grepon, A. (2020). Parkinson’s disease polygenic risk score is not associated with impulse control disorders: A longitudinal study. Parkinsonism and Related Disorders, 75(March), 30–33. https://doi.org/10.1016/j.parkreldis.2020.03.017

Isaias, I. U., Siri, C., Cilia, R., de Gaspari, D., Pezzoli, G., & Antonini, A. (2008). The relationship between impulsivity and impulse control disorders in Parkinson’s disease. Movement Disorders, 23(3), 411–415. https://doi.org/10.1002/mds.21872

Jahanshahi, M., Jenkins, H., Brown, R. G., Marsden, C. D., Passingham, R. E., & Brooks, D. J. (1996). Self-initiated versus externally triggered movements. I. An investigation using measurement of regional cerebral blood flow with PET and movement-related potentials in normal and Parkinson’s disease subjects. Brain, 119(3), 1045–1048. https://doi.org/10.1093/brain/119.3.1045

Joutsa, J., Martikainen, K., Vahlberg, T., & Kaasinen, V. (2012). Effects of dopamine agonist dose and gender on the prognosis of impulse control disorders in Parkinson’s disease. Parkinsonism and Related Disorders, 18(10), 1079–1083. https://doi.org/10.1016/j.parkreldis.2012.06.005

Kelly, M. J., Baig, F., Hu, M. T. M., & Okai, D. (2020). Spectrum of impulse control behaviours in Parkinson’s disease: Pathophysiology and management. In Journal of Neurology, Neurosurgery and Psychiatry (vol. 91, Issue 7, pp. 703–711). BMJ Publishing Group. https://doi.org/10.1136/jnnp-2019-322453

Ko, J. H., Antonelli, F., Monchi, O., Ray, N., Rusjan, P., Houle, S., Lang, A. E., Christopher, L., & Strafella, A. P. (2013). Prefrontal dopaminergic receptor abnormalities and executive functions in Parkinson’s disease. Human Brain Mapping, 34(7), 1591–1604. https://doi.org/10.1002/hbm.22006

Kraemmer, J., Smith, K., Weintraub, D., Guillemot, V., Nalls, M. A., Cormier-Dequaire, F., Moszer, I., Brice, A., Singleton, A. B., & Corvol, J. C. (2016). Clinical-genetic model predicts incident impulse control disorders in Parkinson’s disease. Journal of Neurology, Neurosurgery and Psychiatry, 87(10), 1106–1111. https://doi.org/10.1136/jnnp-2015-312848

Krieger, D. M., Cardoso, S. V., Caumo, W., Valença, G., Weintraub, D., & Rieder, C. R. de M. (2017). Parkinson’s Disease Impulsive-Compulsive Disorders Questionnaire - Current Short (QUIP-CS) - Translation and validation of content of Portuguese Version. Jornal Brasileiro de Psiquiatria, 66(2), 111–115. https://doi.org/10.1590/0047-2085000000158

Lee, J. Y., Lee, E. K., Park, S. S., Lim, J. Y., Kim, H. J., Kim, J. S., & Jeon, B. S. (2009). Association of DRD3 and GRIN2B with Impulse Control and Related Behaviors in Parkinson’s Disease. Movement Disorders, 24(12), 1803–1810. https://doi.org/10.1002/mds.22678

Lee, J. Y., Kim, J. M., Kim, J. W., Cho, J., Lee, W. Y., Kim, H. J., & Jeon, B. S. (2010). Association between the dose of dopaminergic medication and the behavioral disturbances in Parkinson disease. Parkinsonism and Related Disorders, 16(3), 202–207. https://doi.org/10.1016/j.parkreldis.2009.12.002

Leeman, R. F., & Potenza, M. N. (2011). Impulse control disorders in Parkinson’s disease: Clinical characteristics and implications. In Neuropsychiatry (vol. 1, Issue 2, pp. 133–147). https://doi.org/10.2217/npy.11.11

Lepack, A. E., Werner, C. T., Stewart, A. F., Fulton, S. L., Zhong, P., Farrelly, L. A., Smith, A. C. W., Ramakrishnan, A., Lyu, Y., Bastle, R. M., Martin, J. A., Mitra, S., O’Connor, R. M., Wang, Z. J., Molina, H., Turecki, G., Shen, L., Yan, Z., Calipari, E. S., … Maze, I. (2020). Dopaminylation of histone H3 in ventral tegmental area regulates cocaine seeking. Science, 368(6487), 197–201. https://doi.org/10.1126/science.aaw8806

Logan, G. D., Cowan, W. B., & Davis, K. A. (1984). On the ability to inhibit simple and choice reaction time responses: A model and a method. Journal of Experimental Psychology: Human Perception and Performance, 10(2), 276–291. https://doi.org/10.1037/0096-1523.10.2.276

MacDonald, H. J., Stinear, C. M., Ren, A., Coxon, J. P., Kao, J., Macdonald, L., Snow, B., Cramer, S. C., & Byblow, W. D. (2016). Dopamine Gene Profiling to Predict Impulse Control and Effects of Dopamine Agonist Ropinirole. Journal of Cognitive Neuroscience, 28(7), 909–919. https://doi.org/10.1162/jocn

Marín-Lahoz, J., Pagonabarraga, J., Martinez-Horta, S., Fernandez de Bobadilla, R., Pascual-Sedano, B., Pérez-Pérez, J., Gironell, A., & Kulisevsky, J. (2018). Parkinson’s Disease: Impulsivity Does Not Cause Impulse Control Disorders but Boosts Their Severity. Frontiers in Psychiatry, 9. https://doi.org/10.3389/fpsyt.2018.00465

Marković, V., Stanković, I., Petrović, I., Stojković, T., Dragašević-Mišković, N., Radovanović, S., Svetel, M., Stefanova, E., & Kostić, V. (2020). Dynamics of impulsive–compulsive behaviors in early Parkinson’s disease: a prospective study. Journal of Neurology, 267(4), 1127–1136. https://doi.org/10.1007/s00415-019-09692-4

Marques, A., Vidal, T., Pereira, B., Benchetrit, E., Socha, J., Pineau, F., Elbaz, A., Artaud, F., Mangone, G., You, H., Cormier, F., Galitstky, M., Pomies, E., Rascol, O., Derkinderen, P., Weintraub, D., Corvol, J. C., & Durif, F. (2019). French validation of the questionnaire for Impulsive-Compulsive Disorders in Parkinson’s Disease–Rating Scale (QUIP-RS). Parkinsonism and Related Disorders, 63, 117–123. https://doi.org/10.1016/j.parkreldis.2019.02.026

Mata, R., Hau, R., Papassotiropoulos, A., & Hertwig, R. (2012). DAT1 polymorphism is associated with risk taking in the Balloon Analogue Risk Task (BART). PLoS ONE, 7(6). https://doi.org/10.1371/journal.pone.0039135

Martini, A., Ellis, S. J., Grange, J. A., Tamburin, S., Dal Lago, D., Vianello, G., & Edelstyn, N. M. J. (2018). Risky decision-making and affective features of impulse control disorders in Parkinson’s disease. Journal of Neural Transmission, 125(2), 131–143. https://doi.org/10.1007/s00702-017-1807-7

Meder, D., Herz, D. M., Rowe, J. B., Lehéricy, S., & Siebner, H. R. (2019). The role of dopamine in the brain - lessons learned from Parkinson’s disease. In NeuroImage (Vol. 190, pp. 79–93). Academic Press Inc. https://doi.org/10.1016/j.neuroimage.2018.11.021

Nikolova, Y. S., Ferrell, R. E., Manuck, S. B., & Hariri, A. R. (2011). Multilocus genetic profile for dopamine signaling predicts ventral striatum reactivity. Neuropsychopharmacology, 36(9), 1940–1947. https://doi.org/10.1038/npp.2011.82

Papay, K., Mamikonyan, E., Siderowf, A. D., Duda, J. E., Lyons, K. E., Pahwa, R., Driver-Dunckley, E. D., Adler, C. H., & Weintraub, D. (2011). Patient versus informant reporting of ICD symptoms in Parkinson’s disease using the QUIP: Validity and variability. Parkinsonism and Related Disorders, 17(3), 153–155. https://doi.org/10.1016/j.parkreldis.2010.11.015

Pearson-Fuhrhop, K. M., Minton, B., Acevedo, D., Shahbaba, B., & Cramer, S. C. (2013). Genetic Variation in the Human Brain Dopamine System Influences Motor Learning and Its Modulation by L-Dopa. PLoS ONE, 8(4), e61197. https://doi.org/10.1371/journal.pone.0061197

Pearson-Fuhrhop, K. M., Dunn, E. C., Mortero, S., Devan, W. J., Falcone, G. J., Lee, P., Holmes, A. J., Hollinshead, M. O., Roffman, J. L., Smoller, J. W., Rosand, J., & Cramer, S. C. (2014). Dopamine Genetic Risk Score Predicts Depressive Symptoms in Healthy Adults and Adults with Depression. PLoS ONE, 9(5), e93772. https://doi.org/10.1371/journal.pone.0093772

Perez-Lloret, S., Rey, M. V., Fabre, N., Ory, F., Spampinato, U., Brefel-Courbon, C., Montastruc, J. L., & Rascol, O. (2012). Prevalence and pharmacological factors associated with impulse-control disorder symptoms in patients with parkinson disease. Clinical Neuropharmacology, 35(6), 261–265. https://doi.org/10.1097/WNF.0b013e31826e6e6d

Probst, C. C., Winter, L. M., Möller, B., Weber, H., Weintraub, D., Witt, K., Deuschl, G., Katzenschlager, R., & Van Eimeren, T. (2014). Validation of the questionnaire for impulsive-compulsive disorders in Parkinson’s disease (QUIP) and the QUIP-rating scale in a German speaking sample. Journal of Neurology, Neurosurgery and Psychiatry, 261, 936–942. https://doi.org/10.1007/s00415-014-7299-6

Ray, N. J., Brittain, J. S., Holland, P., Joundi, R. A., Stein, J. F., Aziz, T. Z., & Jenkinson, N. (2012). The role of the subthalamic nucleus in response inhibition: Evidence from local field potential recordings in the human subthalamic nucleus. NeuroImage, 60(1), 271–278. https://doi.org/10.1016/j.neuroimage.2011.12.035

Ricciardi, L., Haggard, P., de Boer, L., Sorbera, C., Stenner, M. P., Morgante, F., & Edwards, M. J. (2017). Acting without being in control: Exploring volition in Parkinson’s disease with impulsive compulsive behaviours. Parkinsonism and Related Disorders, 40, 51–57. https://doi.org/10.1016/j.parkreldis.2017.04.011

Ruitenberg, M. F. L., van Wouwe, N. C., Wylie, S. A., & Abrahamse, E. L. (2021). The role of dopamine in action control: Insights from medication effects in Parkinson’s disease. In Neuroscience and Biobehavioral Reviews (Vol. 127, pp. 158–170). Elsevier Ltd. https://doi.org/10.1016/j.neubiorev.2021.04.023

Seeman, P. (2015). Parkinson’s Disease Treatment May Cause Impulse-Control Disorder Via Dopamine D3 Receptors. Synapse, 69(4), 183–189. https://doi.org/10.1002/syn.21805

Sinha, N., Manohar, S., & Husain, M. (2013). Impulsivity and apathy in Parkinson’s disease. Journal of Neuropsychology, 7(2), 255–283. https://doi.org/10.1111/jnp.12013

Smith, K. M., Xie, S. X., & Weintraub, D. (2016). Incident impulse control disorder symptoms and dopamine transporter imaging in Parkinson disease. Journal of Neurology, Neurosurgery and Psychiatry, 87(8), 864–870. https://doi.org/10.1136/jnnp-2015-311827

Takahashi, M., Koh, J., Yorozu, S., Kajimoto, Y., Nakayama, Y., Sakata, M., Yasui, M., Hiwatani, Y., Weintraub, D., & Ito, H. (2022). Validation of the Japanese Version of the Questionnaire for Impulsive-Compulsive Disorders in Parkinson’s Disease-Rating Scale (QUIP-RS). Parkinson’s Disease, 2022. https://doi.org/10.1155/2022/1503167

Trantham-Davidson, H., Neely, L. C., Lavin, A., & Seamans, J. K. (2004). Mechanisms underlying differential D1 versus D2 dopamine receptor regulation of inhibition in prefrontal cortex. Journal of Neuroscience, 24(47), 10652–10659. https://doi.org/10.1523/JNEUROSCI.3179-04.2004

Vaillancourt, D. E., Schonfeld, D., Kwak, Y., Bohnen, N. I., & Seidler, R. (2013). Dopamine overdose hypothesis: Evidence and clinical implications. In Movement Disorders (vol. 28, Issue 14, pp. 1920–1929). https://doi.org/10.1002/mds.25687

Vela, L., Martínez Castrillo, J. C., García Ruiz, P., Gasca-Salas, C., Macías Macías, Y., Pérez Fernández, E., Ybot, I., Lopez Valdés, E., Kurtis, M. M., Posada Rodriguez, I. J., Mata, M., Ruiz Huete, C., Eimil, M., Borrue, C., del Val, J., López-Manzanares, L., Rojo Sebastian, A., & Marasescu, R. (2016). The high prevalence of impulse control behaviors in patients with early-onset Parkinson’s disease: A cross-sectional multicenter study. Journal of the Neurological Sciences, 368, 150–154. https://doi.org/10.1016/j.jns.2016.07.003

Verbruggen, F., Aron, A. R., Band, G. P., Beste, C., Bissett, P. G., Brockett, A. T., Brown, J. W., Chamberlain, S. R., Chambers, C. D., Colonius, H., Colzato, L. S., Corneil, B. D., Coxon, J. P., Dupuis, A., Eagle, D. M., Garavan, H., Greenhouse, I., Heathcote, A., Huster, R. J., … Boehler, C. N. (2019). A consensus guide to capturing the ability to inhibit actions and impulsive behaviors in the stop-signal task. ELife, 8, 1–26. https://doi.org/10.7554/elife.46323

Vriend, C., Nordbeck, A. H., Booij, J., van der Werf, Y. D., Pattij, T., Voorn, P., Raijmakers, P., Foncke, E. M. J., van de Giessen, E., Berendse, H. W., & van den Heuvel, O. A. (2014). Reduced Dopamine Transporter Binding Predates Impulse Control Disorders in Parkinson’s Disease. Movement Disorders, 29(7), 904–911. https://doi.org/10.1002/mds.25886

Vriend, C., Leffa, D. T., Trujillo, J. P., Gerrits, N. J. H. M., de Boer, F. E., Berendse, H. W., van der Werf, Y. D., & van den Heuvel, O. A. (2018). Functional connectivity alterations in Parkinson’s disease during the stop-signal task. BioRxiv, April. https://doi.org/10.1101/304584

Weintraub, D., Siderowf, A. D., Potenza, M. N., Goveas, J., Morales, K. H. D., Duda, J. E., Moberg, P. J., & Stern, M. B. (2006). Dopamine Agonist Use is Associated with Impulse Control Disorders in Parkinson’s Disease. Arch Neurol, 63(7), 969–973. https://www.ncbi.nlm.nih.gov/pmc/articles/PMC1761054/pdf/nihms12138.pdf

Weintraub, D. (2008). Dopamine and Impulse Control Disorders in Parkinson’s Disease. Annals of Neurology, 64(SUPPL. 2), 93–100. https://doi.org/10.1002/ana.21454

Weintraub, D., Stewart, S., Shea, J. A., Lyons, K. E., Pahwa, R., Driver-dunckley, E. D., Adler, C. H., Potenza, M. N., Miyasaki, J., Siderowf, A. D., Duda, J. E., Hurtig, H. I., Colcher, A., Horn, S. S., Stern, M. B., & Voon, V. (2009). Validation of the Questionnaire for Impulsive-CompulsiveDisorders in Parkinson’s Disease (QUIP). Movement Disorders, 24(10), 1461–1467. https://doi.org/10.1002/mds.22571.Validation

Weintraub, D., Koester, J., Potenza, M. N., Siderowf, A. D., Stacy, M., Voon, V., Whetteckey, J., Wunderlich, G. R., & Lang, A. E. (2010). Impulse Control Disorders in Parkinson Disease: a cross-sectional study of 3090 patients. Archives of Neurology, 67(5), 589–595. https://doi.org/10.1001/archneurol.2010.65

Weintraub, D., Mamikonyan, E., Papay, K., Shea, J. A., Xie, S. X., & Siderowf, A. (2012). Questionnaire for impulsive-compulsive disorders in Parkinson’s Disease-Rating Scale. Movement Disorders, 27(2), 242–247. https://doi.org/10.1002/mds.24023

Weintraub, D., & Mamikonyan, E. (2019). Impulse control disorders in Parkinson’s disease. American Journal of Psychiatry, 176(1), 5–11. https://doi.org/10.1176/appi.ajp.2018.18040465

