## Supplemental Tables 1S, 2S, 3S for "Performance on the Balloon Analogue Risk Task and Anticipatory Response Inhibition Task is Associated with Severity of Impulse Control Behaviours in People with Parkinson’s Disease"

**Table 1S.** Occurrence of polymorphisms for dopamine genetic risk score

| | DRD1 rs4532 | | | DRD2 rs1800497 | | | DRD3 rs6280 | | | COMT rs4680 | | | DAT rs28363170 | | |
|---|---|---|---|---|---|---|---|---|---|---|---|---|---|---|---|
| | A/A | A/G | G/G | C/C | C/T | T/T | T/T | C/T | C/C | G/G | G/A | A/A | 9/9 | 9/10 | 10/10 |
| Score | 0 | 1 | 1 | 1 | 0 | 0 | 0 | 1 | 1 | 0 | 1 | 1 | 1 | 1 | 0 |
| Predict freq | 0.39 | 0.47 | 0.14 | 0.58 | 0.36 | 0.06 | 0.36 | 0.48 | 0.15 | 0.17 | 0.49 | 0.35 | 0.09 | 0.42 | 0.49 |
| Actual freq | 0.36 | 0.52 | 0.18 | 0.56 | 0.39 | 0.04 | 0.36 | 0.50 | 0.14 | 0.18 | 0.47 | 0.35 | 0.10 | 0.39 | 0.49 |

DRD1: dopamine receptor D1; DRD2: dopamine receptor D2; DRD3: dopamine receptor D3; COMT: catechol-O-methyltransferase; DAT: dopamine transporter. A: adenine; G: guanine; C: cytosine; T: thymine. Predict freq: expected mutation frequency in population. Actual freq: observed frequency in current population.

**Table 2S.** Univariate linear regression analysis of variables associated with the frequency of impulse control behaviours.

| **ICB (n = 23) no ICB (n = 27)** | | | | |
|---|---|---|---|---|
| | **β** | **SE** | **p value** | **95 % CI (β)** |
| Average collection pumps | -0.005 | 0.21 | .980 | [-0.43, 0.42] |
| Age | 0.18 | 0.26 | .490 | [-0.35, 0.71] |
| DGRS low | -0.04 | 5.11 | .994 | [-10.3, 10.2] |
| **Gender (male)** | **9.76** | **4.47** | **.034** | **[0.77, 18.8]** |
| LEDD DA | -0.006 | 0.02 | .765 | [-0.05, 0.04] |
| LEDD Total | 0.004 | 0.006 | .513 | [-0.008, 0.02] |
| Negative Reinforcement | 6.92 | 4.77 | .153 | [-2.67, 16.5] |
| Positive Reinforcement | -5.24 | 5.71 | .363 | [-16.7, 6.25] |
| SSRT stop both | 0.03 | 0.03 | .400 | [-0.04, 0.10] |
| **UPDRS I&II** | **0.88** | **0.19** | **<.001** | **[0.50, 1.27]** |
| **Years on DA** | **1.40** | **0.43** | **.002** | **[0.52, 2.27]** |
| **Years since diagnosis** | **1.10** | **0.32** | **.001** | **[0.44, 1.75]** |

Response variable: score on Questionnaire for Impulsive-Compulsive Disorders in Parkinson's Disease rating scale. ICB: impulse control behaviour (n: number); DGRS: dopamine genetic risk score; LEDD: levodopa equivalent daily dose; DA: Dopamine Agonist; SSRT: stop signal reaction time; UPDRS: Unified Parkinson's Disease Rating Scale; β: coefficient, SE: standard error, CI: confidence interval. Significant values in bold ($p < .05$).

**Table 3S. Continuous Independent variable collinearity for the Dopamine Agonist group.**

| *Coefficient* | **Years since diagnosis** | **Years on DA** | **DA LEDD** | **UPDRS** | **SSRT both** | **Negative Reinforcement** |
|---|---|---|---|---|---|---|
| **Years since diagnosis** | | **0.85 (<.001)** | 0.12 (.420) | **0.65 (<.001)** | -0.18 (.224) | -0.07 (.647) |
| **Years on DA** | **0.85 (<.001)** | | 0.26 (.065) | **0.55 (<.001)** | -0.14 (.330) | -0.24 (.099) |
| **DA LEDD** | 0.12 (.420) | 0.26 (.065) | | -0.03 (.837) | -0.24 (.090) | -0.28 (.051) |
| **UPDRS** | **0.65 (<.001)** | **0.55 (<.001)** | -0.03 (.837) | | 0.08 (.559) | 0.12 (.408) |
| **SSRT both** | -0.18 (.224) | -0.14 (.330) | -0.24 (.090) | 0.08 (.559) | | -0.13 (.354) |
| **Negative Reinforcement** | -0.07 (.647) | -0.24 (.099) | -0.28 (.051) | 0.12 (.408) | -0.13 (.354) | |

Correlation coefficient (p value). LEDD: levodopa equivalent daily dose; DA: dopamine agonist; UPDRS: Unified Parkinson's Disease Rating Scale;

Significant values in bold ($p < .05$).
